# Trap and kill of environmental microbes: Validation of a novel decontamination technology in Hospital ICU setting

**DOI:** 10.1101/2020.08.02.20166801

**Authors:** Sanju Jose, Kruttika S. Phadke, Janani Venkatraman, Bhuvana Krishna, Sriram Sampath, Santanu Datta, Savitha Nagaraj, Arindam Ghatak

**Affiliations:** St. John’s Hospital and Medical College, Bangalore, Karnataka, India; Biomoneta Research Private Limited, Bangalore, Karnataka, India; Bugworks Research India, Bangalore, Karnataka, India

## Abstract

Nosocomial infections, also known as hospital-acquired infections (HAI), appear 48 hours or more after hospital admission and are independent of the original infirmity of the patient. To prevent or to reduce HAI, the central paradigm is to construct protective barriers between the large number of people who are sick and whose immune systems are compromised in the precincts of the hospital. Microbes that result in HAI do so by two routes of infection: touch and aerosol. We describe here ZeBox technology, a voltage induced synergistic killing of the microbe on designed surfaces, as a game-changer in this domain. Its kill rate is hitherto unmatched by any known chemical or non-chemical (viz; UV, ionisation) technology. In an enclosed test chamber, under challenge conditions, ZeBox technology can kill about a billion microbes in 10 minutes. When tested under clinical settings, the device could effectively reduce microbes, both from air and surfaces with more than 90% efficiency. The optimum requirement to reduce HAI would be to construct an online microbicidal device that operates in a continuous trap and kill mode in the background of people and patient movement, and decontaminates air and surfaces. We present unequivocal data to fortify our claims of online, continuous, safe, trap and kill mechanism of ZeBox technology.

## Introduction

Healthcare-associated infections (HAIs) are a growing concern for clinical practice worldwide. Close to 2 million patients contract HAIs in the U.S. every year, out of which nearly 100,000 patients die. The overall HAI rate is about three-fold higher in the developing world, with the risk of contracting device associated infections being as much as 15-19 fold higher. (1) These infections are often due to multidrug-resistant (MDR) bacteria, which have increasingly few treatment options (2). Additionally, the risk of contracting HAIs increases if the prior bed or room occupant suffered from infection; this risk is as much as four-fold higher for an Acinetobacter infection.

Pathogenic microorganisms persist in the environment despite regular cleaning protocols instituted by healthcare spaces. *Acinetobacter* sp. and *Clostridium difficile* can persist for as much as five months while *Pseudomonas* can linger on surfaces for a long as 16 months. Microbial particles can be deposited on surfaces distant from the source of contamination via airborne dispersion and can be further dispersed via contact with healthcare workers and cross-contamination. (3,4)

Microbial contamination in hospital wards is concentrated in hard-to-reach surfaces such as the floor under beds and bed wheels as compared to higher levels of a room. This correlates both with the source of infection (patients in beds) and the fact that air trapped under beds and instruments is not efficiently cycled through wall mounted air purification units. There is a pressing need to design microbial decontamination devices that function near microbial reservoirs.

Infection control in most hospitals today focus on surface disinfection and hand hygiene, which are dependent on staff efficiency and compliance. It is an accepted fact that systems that are dependent on human behaviour suffer from inefficiencies due to human fatigue. In fact, there is evidence to show that thoroughness of cleaning drops by 10-20% after the education and supervision period ends and that there is inconsistent adherence to room cleaning and hand hygiene protocols in the absence of a threat of an epidemic. (5)

Technologies that lower the environmental load in healthcare systems with the goal of reducing the rate of HAIs have been explored in the U.S and EU over the last 5-10 years. Most of these technologies have been either light-based (Xenon-powered laser producing robots by Xenex; blue-white LED fixtures by Indigo Clean) or filter based (Cleanroom H13 by IQAir). Clinical data generated by especially Xenex suggests that reducing pathogen load in the environment can lower the rate of infections in hospitals. E.g., one study showed that nightly pulsed-xenon ultraviolet disinfection working with dedicated housekeeping staff reduced SSIs in Class I procedures by 46%. (6,7)

However, application of UV based technologies for room (ICUs, wards) decontamination is not compatible with the patient load seen in most hospitals in India, where rooms are rarely empty, and UV radiation cannot be used in the presence of humans. Filter based technologies suffer from problems of clogging and hence reduced efficiency. Additionally, medical HEPA filters are expensive to buy and to utilize.

To this end, we have validated an environmental decontamination device, powered by a novel decontamination technology, ZeBox Technology. The decontamination devices powered by ZeBox Technology can extract and kill microorganisms from the air and nearby surfaces [Cite patent]. This device can function in the presence of patients and hospital staff and needs little staff training or intervention. It acts near microbial reservoirs, namely the zone in and around a patient bed, and aims to create a virtual near-sterile envelope around a patient bed.

### ZeBox Technology and Device description

The technology innovation is called the ZeBox, where Ze stands for Zeta Potential, the potential or charge difference across microbial surfaces. The movement of highly charged particles like microbes, can be manipulated by the application of electric fields. We apply directed electric fields across the flow of air bearing microbial particles to accelerate their trapping and retention on defined surfaces. Additionally, the field potentiates said surface, rendering it microbicidal in nature. Designing the flow of air to be parallel to the antimicrobial surface significantly eliminates clogging and masking of the treated surface by minimizing build-up of dust and microbial debris. Clogging is a major problem with traditional filter based devices, where filters are placed perpendicular to air flow. Devices, powered by ZeBox Technology, extract and kill microbes from air and nearby surfaces by synergistically combining three disparate aspects - defined chemically treated surface that can be rendered microbicidal, defined electric field to trap and retain microbes on the surface for the duration required for kill, and airflow design to maximize impact of microbes on the surface with minimal clogging.

The device tested in the present study is 66cm x 35cm x 20 cm in dimension, mounted on a stand with a footprint of 40cm x 45cm. The device uses air as a carrier medium to pull in microbes from air and nearby surfaces, which then passes between channels created by surfaces with specialized architecture. These surfaces get activated when a modulated electric field is applied. This electric field also moves the microbes and impels them onto these activated surfaces, where they are lysed. The device has an airflow rate of 150CFM.

## Material and Methods

### 1. Challenge tests

#### A. Test setup

An air-sealed test chamber of 1000Liters air volume was built with multiple sampling ports and nebulization port. The environmental parameters viz relative humidity and temperature could be monitored using a probe located inside the chamber. During experiments, various microorganisms were aerosolized using a 6-jet collision nebulizer into this chamber, and device efficiency was monitored by collecting and measuring microbial concentration at different time intervals.

#### B. Aerosolization of test microbes

A 6-jet Collison nebulizer (MESA LABS, BGI) was used to aerosolize the test microbes into the test chamber. Dry air from a compressed air cylinder was used at a pressure of 10 lb/in2 to operate the nebulizer. The nebulizer produces bioaerosols of a 2-5µm diameter that allows them to float in the air present in the test chamber for a definite period. The length of the nebulization period varied depending on the type of experiment and microorganism.

#### C. Sampling of air for viable microbes

The airborne survival of the test microbe and the activity of the air decontamination devices were determined by collecting the air from the chamber at the rate of 30liter/min using a customized collection chamber filled with sterile buffer (1x Phosphate buffer saline, pH 7.2). Collected samples were analyzed to understand the quantity of viable microorganism present by diluting and plating them onto suitable growth media. The plated samples were incubated at 37± 2 °C for bacteria and 25± 2 °C for fungal species and allowed to grow for 24 hours, individual colonies were enumerated, and the final concentration of the microbial load was calculated thereafter.

#### D. Cultivation of test microorganisms

To validate the efficiency of the decontamination device, *Escherichia coli, Pseudomonas aeruginosa, Staphylococcus aureus, Candida albicans, Aspergillus fumigatus, Mycobacterium smegmatis* were used. For growing *Escherichia coli, Pseudomonas aeruginosa* and *Staphylococcus aureus*, Luria Bertani broth were used. For growing *Candida albicans* Potato dextrose broth was used, while for *M. smegmatis* Middlebrook 7H9 broth was used. For enumeration of E.coli, samples were plated on Luria Bertani agar; Cetrimide agar was used as a selective for the growth and isolation of Pseudomonas aeruginosa. Cetrimide inhibits the growth of many microorganisms while allowing Pseudomonas aeruginosa to develop typical colonies. Cetrimide also enhances the production of Pseudomonas sp. pigments, e.g. pyocyanin which shows a characteristic blue-green colour. For quantification of Staphylococcus, Mannitol Salt Agar plates were used. Mannitol Salt is used as a selective and differential media for the growth of Staphylococcus sp. Staphylococcus sp. can tolerate the high salt concentration (7.5% of NaCl) and ferment Mannitol present in the media. Phenol Red is used as an indicator.

Candida and Aspergillus spores were enumerated using Rose-Bengal Chloramphenicol Agar plates. This media is supplemented with Chloramphenicol, which suppresses bacterial growth. Rose-Bengal controls the size and height of the mould population, thereby assisting in a proper enumeration. It also prevents the overgrowth of moulds.

### 2. Testing device efficiency in Hospital ICU

#### A. Test Setup

The study was conducted in a single bed ICU as recommended by the the Hospital Internal Ethics Committee. The room was mechanically ventilated with filtered and tempered air at 22.6±1.9°C with no humidification. Domestic and nursing staff share routine cleaning; near-patient sites are cleaned by nurses twice daily at 7 am and 7 pm. Cleaning is detergent-based, using wipes (Vernacare Tuffie™ wipes) and detergent (Hospec™) for general surfaces. Terminal cleaning of the bed-space is performed following discharge. Two positions inside the ICU was chosen to be monitored during the entire course of study. Air and surface samples were collected from these mentioned positions (n= 240 for air samples and n=260 for surface samples). Samples were collected between 2-3 pm, 7-8 hours post-cleaning of near-surfaces. The surface samples indicate the rate at which microorganisms deposits onto these designated surfaces.

#### B. Air Sample collection

A Handheld air sampler (SAS Super 100) was used, which could sample 100lts of air per minute (8). Tryptic Soy Agar and Sabouraud dextrose agar plates were used to sample bacteria and fungi, respectively from the air. A fixed volume of air was sampled using the bio-sampler. Plates were placed in and removed from the bio-sampler in an aseptic manner. Plates were incubated at 25±2° C (for fungal cultivation) and 37±2° C (for bacterial cultivation) for 48 hours. Post-incubation, the number of colonies appeared were enumerated and converted to CFU/m^3^ using statistical conversion provided by the manufacturer. Control plates were used to ensure the sterility of the entire process.

#### C. Surface Sample collection

A cotton swab was moistened with sterile phosphate-buffered saline (1X PBS; pH 7.2) solution using aseptic technique to prevent cross-contamination and was used to wipe a surface of 100cm^2^ area as mentioned in CDC Guideline (EMERGENCY RESPONSE RESOURCES https://www.cdc.gov/niosh/topics/emres/unp-envsamp.html). The sampled swab was placed in a sterile conical vial containing 1ml of sterile phosphate-buffered saline (PBS) solution. The entire 1ml solution was then plated on to Tryptic Soy Agar and Sabouraud dextrose agar plates for quantification of bacteria and fungi, respectively. Plates were incubated at 25°C (for fungal cultivation) and 37°C (for bacterial cultivation) for 48 hours.

Post-incubation, the number of colonies that appeared were enumerated. Control plates were used to ensure the sterility of the entire process.

## Results

### 1. Validation of decontamination device under challenge condition

The experimental strategy consists of the evaluation of the capability of a decontamination device powered by our proprietary ZeBox Technology to decontaminate a confined space of microorganisms. A broad spectrum of microorganisms was tested in this study, including Gram-positive, Gram-negative, *Mycobacterium smegmatis*, Yeast and Spores of molds. The starting microbial load varied for microorganisms, ranging from 5log_10_ to 10log_10_. The device could eliminate the high concentration of microbial load floating in the air in less than 10 minutes (Table 01, Fig 04). The devices could reduce 9.9log_10_ E.coli in 10 minutes, which could be written alternately as 99.999999999% reduction. For microorganism like *Staphylococcus aureus, Pseudomonas aeruginosa, Candida albicans, Aspergillus fumigatus* spores and *Mycobacterium smegmatis* the device could reduce 5log_10_ to 9log_10_ or 99.999-99.9999999% of viable microbial load based on the starting concentration.

**Table 01:** Comparing efficiency of the decontamination device tested against a broad range of microorganisms. A time-course experiment chart signifies that under a challenged environmental chamber, the efficiency to eliminate airborne microbes increases significantly between 7 minutes to 10 minutes. The number of viable microorganisms aerosolized were more than million in all cases, thus a “log_10_” scale was used to decipher the elimination capability of the device.

|  | Reduction of microbial load using ZeBox (log<sub>10</sub> Scale) |  |  |  |
| --- | --- | --- | --- | --- |
|  | 3 mins<br>of<br>runni<br>ng<br>ZeBox<br>power<br>ed<br>device | 5 mins<br>of<br>runni<br>ng<br>ZeBox<br>power<br>ed<br>device | 7 mins<br>of<br>runni<br>ng<br>ZeBox<br>power<br>ed<br>device | 10<br>mins<br>of<br>runni<br>ng<br>ZeBox<br>power<br>ed<br>device |
| *Escherichia coli* | 0.46 | 1.67 | 3.16 | 9.92 |
| *Staphylococcus aureus* | 0.86 | 1.46 | 2.62 | 5.56 |
| *Pseudomonas aeruginosa* | 0.28 | 1.22 | 4.58 | 7.90 |
| *Candida albicans* | 0.56 | 1.48 | 2.18 | 6.59 |
| *Aspergillus fumigatus*<br>spores | 0.68 | 1.14 | 1.38 | 5.27 |
| *Mycobacterium smegmatis* | 0.46 | 2.19 | 3.44 | 6.68 |

**Figure 01:**
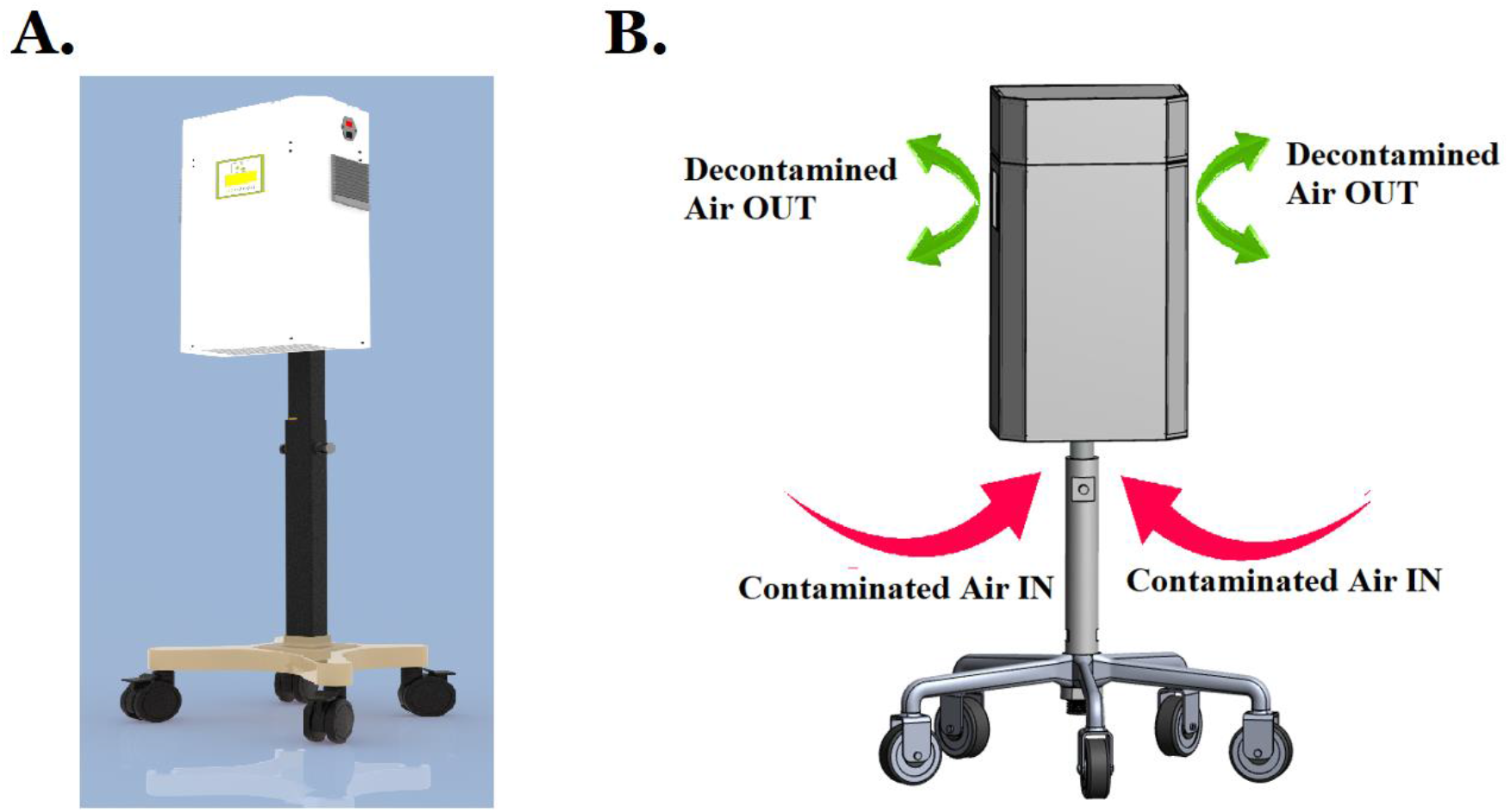
A. Rendered image of the device that was used for validation in laboratory and Hospital ICU. B. Graphical representation of the device used. The device pulls in contaminated air from the bottom, and releases decontaminated air from two sides. The Contaminated air passes through an array of specialized surfaces, which in the context of the device, trap and kill microbes.

**Figure 02:**
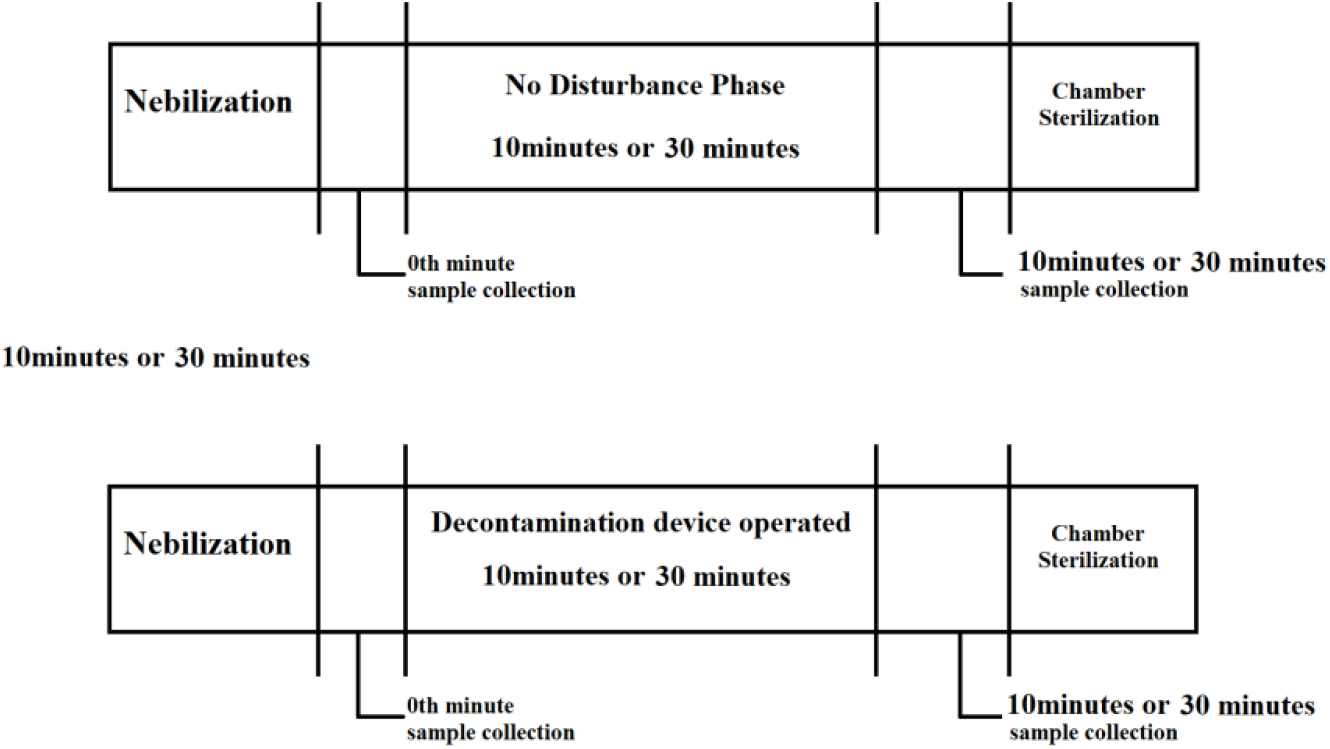
Schematics for the challenge environment experiments, performed to validate the ZeBox Technology powered devices. The aerosolization process was carried out using a 6-jet nebulizer, while air samples were collected from the test chamber using customized glasswares. Collected samples were quantified using standard enumeration method.

**Figure 03.**
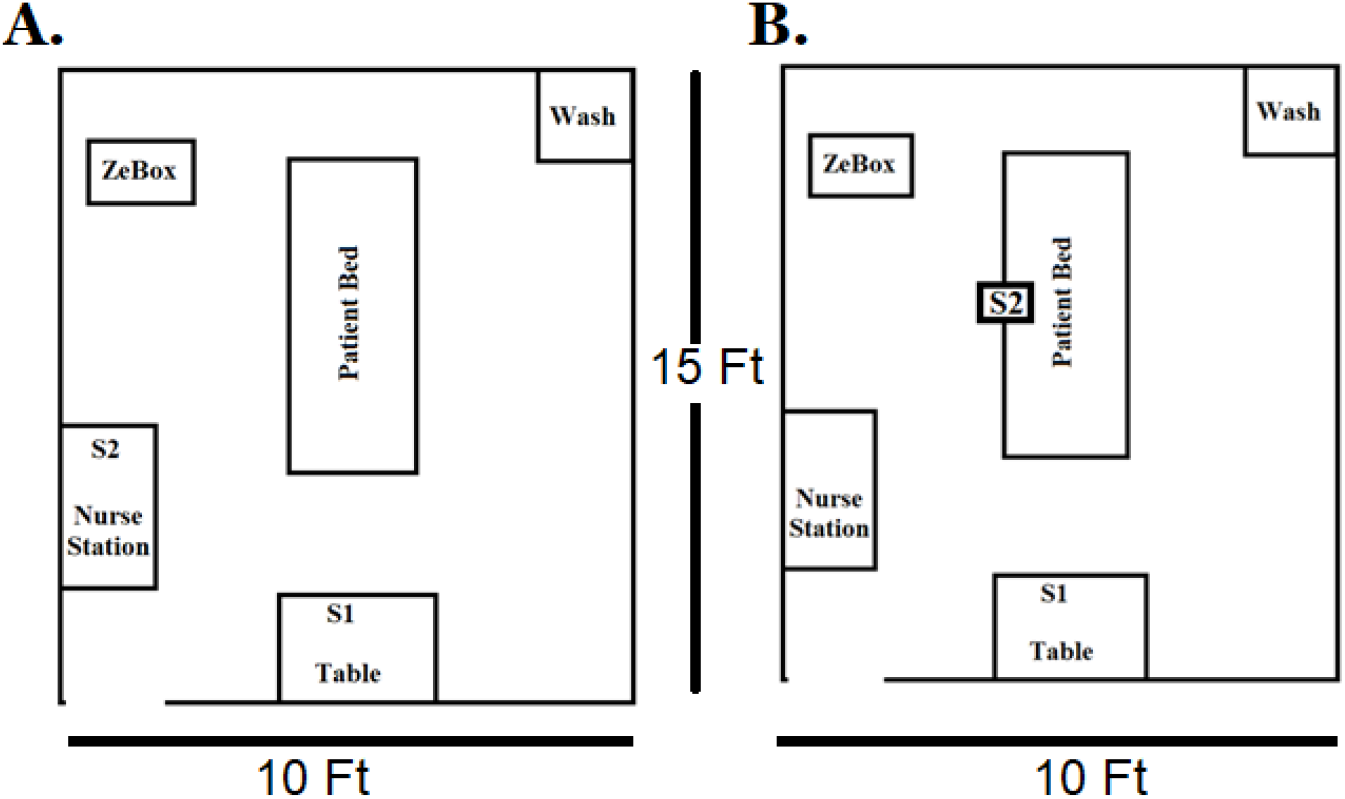
A. ICU Room schematics for collecting air samples. The room has a dimension of 15×10 Ft, samples were collected from positions S1 and S2 for quantification of total bacterial and fungal population. Position S1 and S2 were 10 Feet and 6 Feet away from the deployed device respectively. B. ICU Room schematics for collecting surface samples. The room has similar dimensions as mentioned earlier, samples were collected from Positions S1 (medicine and reporting table) and S2 (bed rails) as recommended in the CDC protocols. Sample positions S1 and S2 were 10 feet and 4 feet away from the deployed device

**Figure 04:**
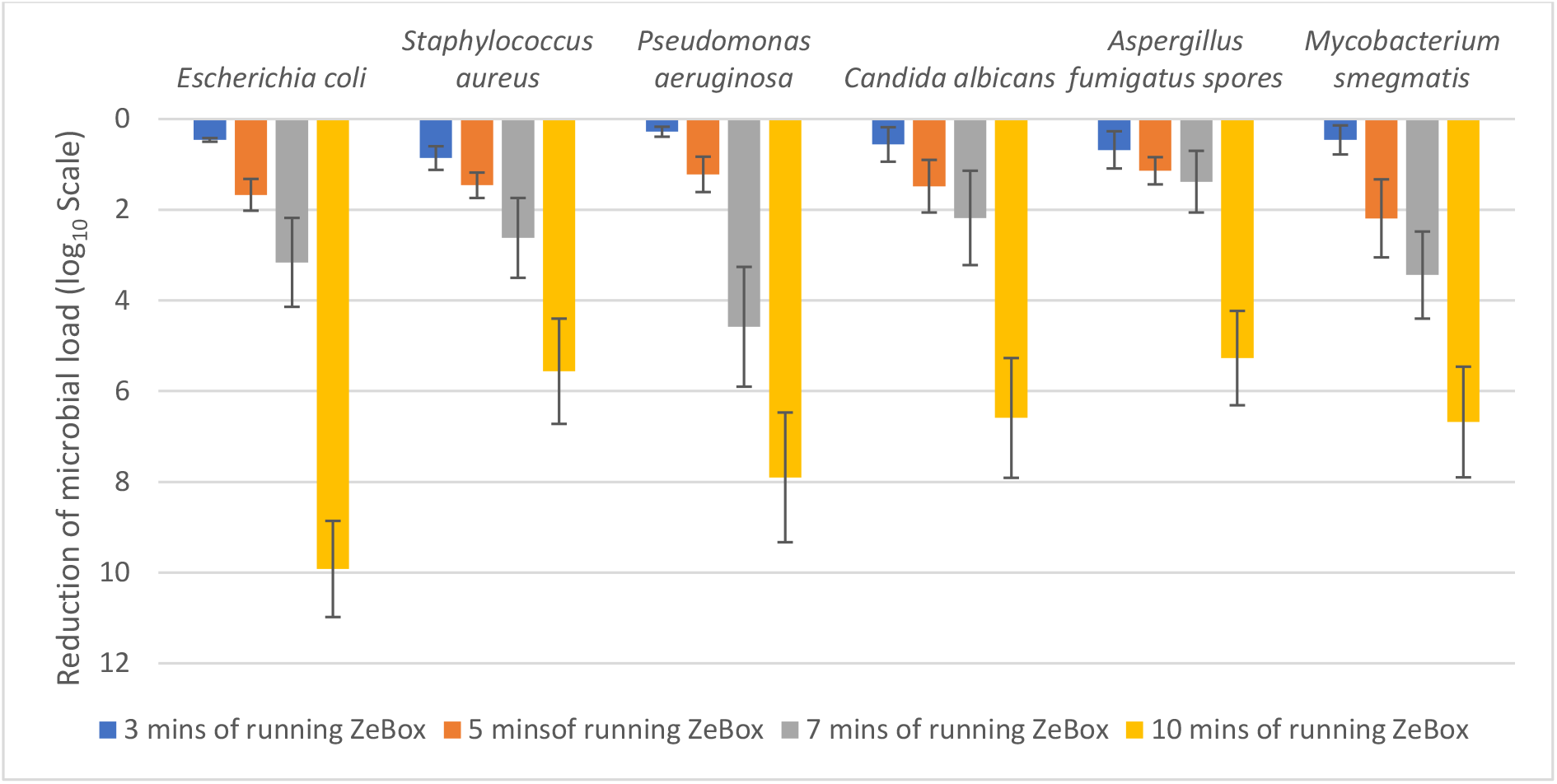
Validation of the decontamination device in a test chamber. Aerosolized microorganisms were used to validate the efficiency of the device under challenge conditions. The device could eliminate wide array of microorganisms, the efficiency primarily depending on the microbial load that could be generated during the process.

### 2. Validation in Hospital ICU

#### A. Airborne microorganisms

The environmental microbial load was monitored in a single ICU bed room with patients and regular hospital functionalities. Air samples were collected as mentioned previously and were estimated for total culturable microbial load. Baseline samples were collected over a period of two months to understand the microbial distribution present in the room, followed by device deployment and sample collection. The bacterial load in the air before deployment were ranging from 2000-2600 CFU/m^3^ while the fungal load ranged from 150-250 CFU/m^3^. Post-deployment of the decontamination device, the microbial load was reduced to 60-100 CFU/m^3^ and 8-17 CFU/m^3^ for bacterial and fungal population, respectively (Table 02, Figure 5)

**Table 02:** Air Samples were from Position 01 and Position 02 to enumerate the total bacterial and fungal load. A total of 240 samples were collected and analysed for bacterial and fungal counts. When the decontamination device powered by ZeBox Technology was deployed, microbial load was reduced by 90-95% for bacteria and 75-80% for fungal population.

|  | Bacterial Load (CFU/m<sup>3</sup>) |  | Fungal Load (CFU/m<sup>3</sup>) |  |
| --- | --- | --- | --- | --- |
|  | Position 01 | Position 02 | Position 01 | Position 02 |
| Pre-deployment | 2397 | 2351 | 173 | 196 |
| Post-deployment | 81 | 74 | 49 | 45 |

**Table 03:** Surface Samples were collected from Position 01 and Position 02 to enumerate the total bacterial and fungal load. A total of 260 samples were collected and analysed for bacterial and fungal count. When the decontamination device powered by ZeBox Technology was deployed, microbial load was reduced by 80-90%% for bacterial and 75-84% for fungal population.

|  | **Bacterial Load (CFU/100cm<sup>2</sup>)** |  | **Fungal Load (CFU/100m<sup>2</sup>)** |  |
| --- | --- | --- | --- | --- |
|  | **Position 01** | **Position 02** | **Position 01** | **Position 02** |
| **Pre-deployment** | 138 | 36 | 7 | 6 |
| **Post-deployment** | 10 | 9 | 1 | 1 |

**Table 04:** Correlation between various enumeration scales. “log_10_” scale is used to describe reduction in microbial load by the device under challenge conditions, “Percentage” scale is used to describe reduction in microbial load in clinical trials.

| Reduction |  |  |
| --- | --- | --- |
| log<sub>10</sub> scale | Percentage (%) Scale | Fold Scale |
| 1 | 90 | 10 |
| 2 | 99 | 100 |
| 3 | 99.9 | 1000 |
| 4 | 99.99 | 10000 |
| 5 | 99.999 | 100000 |

**Figure 5:**
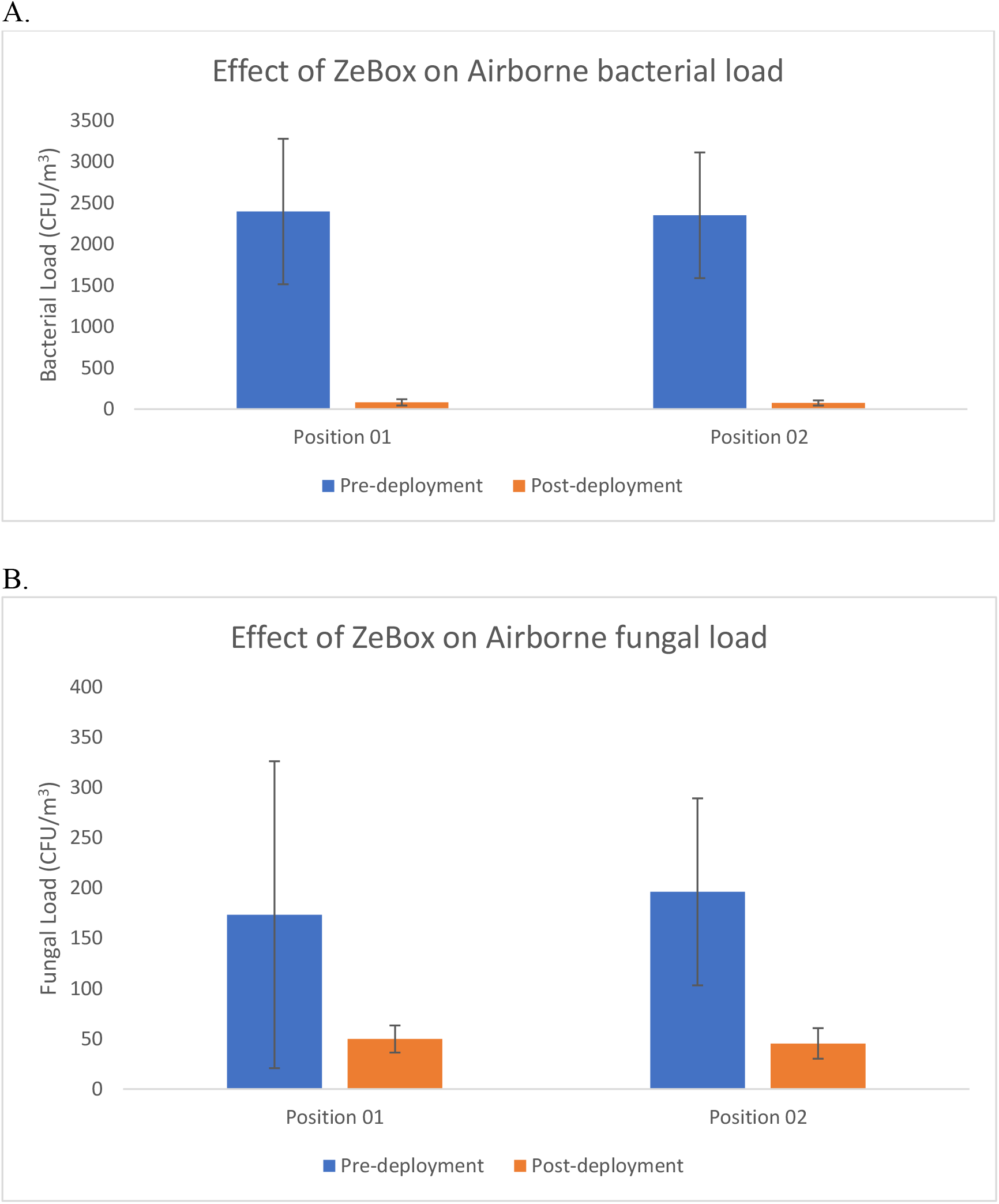
Position S1 and S2 were selected for analysis of air samples. Samples were collected using standard methods as described previously. Prior to device deployment, the bacterial load of the room varied between 2000-2600 CFU/m^3^ while the fungal load ranged from 150-250 CFU/m^3^. Post-deployment of the device, the microbial load was reduced to 60-100 CFU/m^3^ and 8-17 CFU/m^3^ for bacterial and fungal population, respectively.

#### B. Surface borne microorganisms

The surface microbial load was monitored using techniques mentioned previously. Samples were collected from two different positions in a single ICU bed cubicle. Patient bed rail and a medicine storage table were selected as sampling locations. Microbial load, especially bacterial, were found to be high in the medicine table as compared to the other position in the baseline samples. Bacterial load before deploying the decontamination device ranged from 81-143 CFU/100cm^2^ and 26-43 CFU/100cm^2^ on patient bed rails and medicine table respectively. Fungal load before deploying the decontamination device ranged from 6-8CFU/100cm^2^ and 4-7CFU/100cm^2^ on patient bed rails and medicine table, respectively. Post-deployment of the decontamination device, the bacterial load was reduced to 8-12 CFU/100cm^2^ and 6-12CFU/100cm^2^ on patient bed rails and medicine table respectively, while the fungal load was reduced to 1-2 CFU/100cm^2^ and 1-2 CFU/100cm^2^ on patient bed rails and medicine table respectively (Table 02, Figure 6)

**Figure 6:**
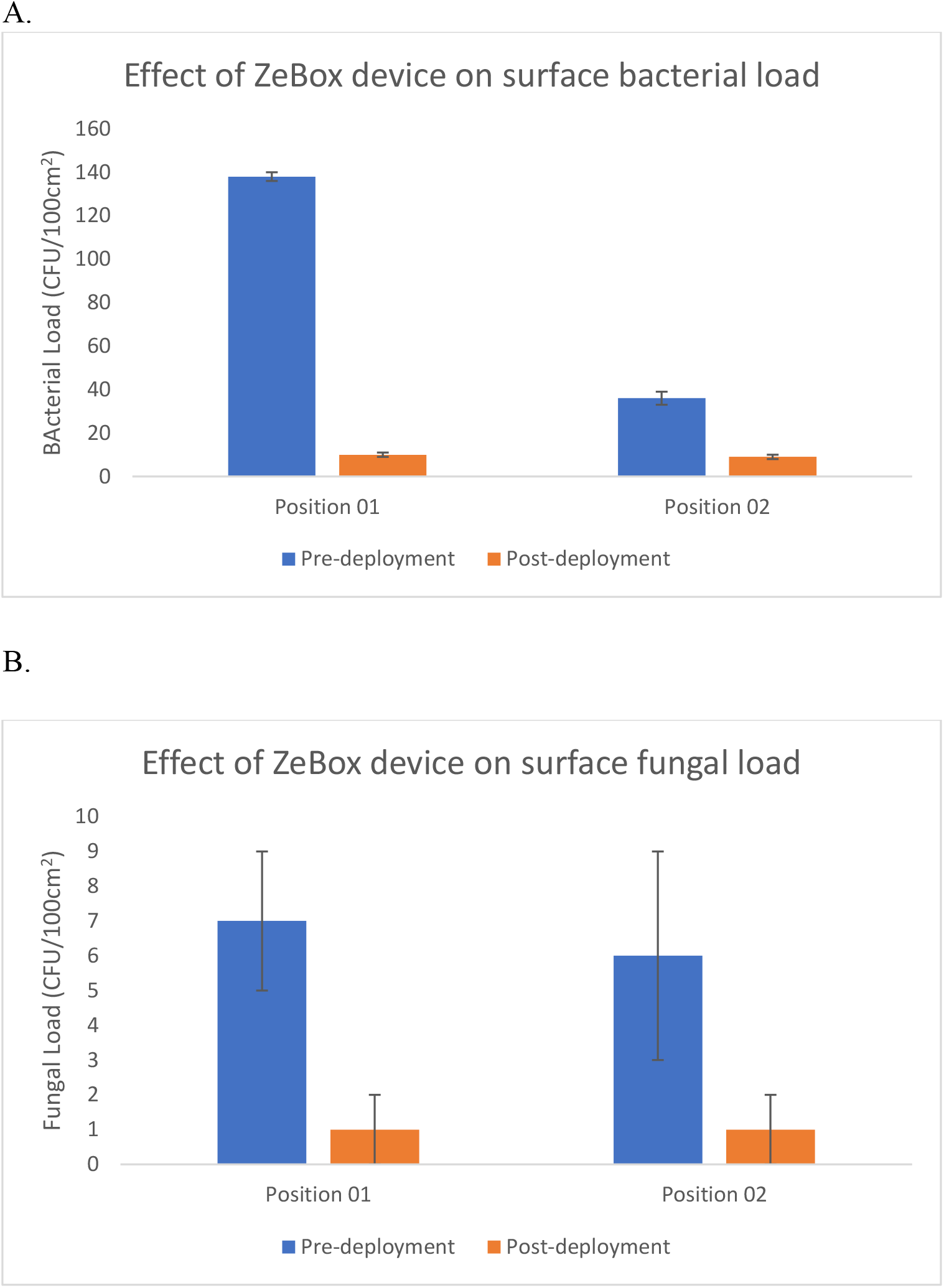
Position S1 and S2 were selected for analysis of surface samples. Samples were collected using standard methods as described previously. 81-143 CFU/100cm^2^ and 26-43 CFU/100cm^2^ on patient bed rails and medicine table respectively. Fungal load before deploying the decontamination device ranged from 6-8CFU/100cm^2^ and 4-7CFU/100cm^2^ on patient bed rails and medicine table, respectively. Post-deployment of the decontamination device, the bacterial load was reduced to 8-12 CFU/100cm^2^ and 6-12CFU/100cm^2^ on patient bed rails and medicine table respectively, while the fungal load was reduced to 1-2 CFU/100cm^2^ and 1-2 CFU/100cm^2^ on patient bed rails and medicine table respectively.

### 3. Significance of using various quantification scale for analysis

In the present article, we have used three different quantification scale to analyse the data generated. For validation of the device and technology under challenge environments, we have used “log_10_” scale, while in clinical settings we have used “Percentage (%)”. At times, for comparative analysis, the “fold” difference was used. Each scale is appropriately used to best emphasize device superiority.

## Discussion

The decontamination devices powered by the novel ZeBox Technology show extreme efficiency in reducing airborne microbial load under challenge conditions, reducing 5log_10_-9log_10_ depending on the test microorganism. The devices are also capable of eliminating various types of microorganisms, viz bacteria, fungi and their spores, floating in the air. Validation of devices in clinical environment resulted in a 0.8log_10_-2log_10_ reduction in hospital ICU environmental microbial load, both bacteria and fungi. Initial analysis was performed with air samples, which were further expanded to surface samples. Successful deployment in a hospital environment reduced the microbial load (bacteria and fungi) from both air and surfaces. The devices tested in the clinical environment underwent IEC 60601-1 and IEC60601-1-2 before deployment. These certifications ensured that the decontamination devices are electrically safe to be used alongside patients in a clinical environment.

## Conclusion

Effective decontamination technology that aids infection control in healthcare spaces must

1. kill pathogenic or contaminating microbes instead of merely trapping,
2. operate continuously and safely in human presence, and
3. require near-zero manual intervention while operating close to the source of infection or contamination.

No other technology being evaluated globally meets all these requirements. While filtration technologies fail to meet the first criterion, UV and ionization based technologies fail to meet the last two. The present disclosed unique, extremely effective, energy-efficient technology, ZeBox satisfies all these attributes.

The devices powered by patented ZeBox technology effectively eliminated microorganism like *Staphylococcus aureus, Pseudomonas aeruginosa, Candida albicans, Aspergillus fumigatus* spores and *Mycobacterium smegmatis*. The device reduced 5log_10_ to 9log_10_ or 99.999-99.9999999% of viable microbial load based on the starting concentration. This novel decontamination technology effectively eliminated the microbial population present in an environment with efficiency close to 90%, both from the air as well as high contact surfaces like patient bed rails. Reducing the environmental microbial load will reduce the occurrence of nosocomial infections in healthcare environments.

Though we successfully show the device’s capability in eliminating bacterial and fungal load from the environment, further study needs to be done with viruses, especially respiratory viruses. Nevertheless, this study successfully evaluates a novel decontamination technology that could be used not only in hospitals but also in other areas where maintaining low-bioburden is a challenge.

## Data Availability

All data is presented in the manuscript

## Acknowledgement

AG and JV deeply acknowledge the contribution of Mr. Ramanujan KS of Biomoneta Research for his technical and intellectual support. We acknowledge staff members Department of Microbiology and Department of Emergency Medicine St.John’s Medical College and Hospital, Bangalore for their co-operation during this study.

## Author Contribution

JV,SD and AG conceptualized and developed the technology, challenge test methods. KSP executed the challenge test under the supervision of JV and AG. BK,SS and SN designed the clinical environment study, SJ performed the clinical environment study under the supervision of BK and SN. JV and AG managed the funding.

## References

1. Allegranzi B, Bagheri Nejad S, Combescure C, et al. Burden of endemic health-care-associated infection in developing countries: systematic review and meta-aanalysis. Lancet. 2011;377(9761):228–241. doi:10.1016/S0140-6736(10)61458-4

2. Nseir S, Blazejewski C, Lubret R, Wallet F, Courcol R, Durocher A. Risk of acquiring multidrug-resistant Gram-negative bacilli from prior room occupants in the intensive care unit. Clin Microbiol Infect. 2011;17(8):1201–1208. doi:10.1111/j.1469-0691.2010.03420.x

3. Kramer A, Schwebke I, Kampf G. How long do nosocomial pathogens persist on inanimate surfaces? A systematic review. BMC Infect Dis. 2006;6:130. Published 2006 Aug 16. doi:10.1186/1471-2334-6-130

4. King, M., Noakes, C., Sleigh, P. and Camargo-Valero, M., 2013. Bioaerosol deposition in single and two-bed hospital rooms: A numerical and experimental study. Building and Environment, 59, pp. 436–447.

5. Peterson, K., Novak, D., Stradtman, L., Wilson, D., & Couzens, L. (2015). Hospital respiratory protection practices in 6 U.S. states: a public health evaluation study. American journal of infection control, 43(1), 63–71. doi: 10.1016/j.ajic.2014.10.008

6. Catalanotti A, Abbe D, Simmons S, Stibich M. Influence of pulsed-xenon ultraviolet light-based environmental disinfection on surgical site infections. Am J Infect Control. 2016;44(6):e99–e101. doi:10.1016/j.ajic.2015.12.018

7. Stibich M, Stachowiak J, Tanner B, et al. Evaluation of a pulsed-xenon ultraviolet room disinfection device for impact on hospital operations and microbial reduction. Infect Control Hosp Epidemiol. 2011;32(3):286–288. doi:10.1086/658329

8. Napoli, C., Marcotrigiano, V. & Montagna, M.T. Air sampling procedures to evaluate microbial contamination: a comparison between active and passive methods in operating theatres. BMC Public Health 12, 594 (2012). https://doi.org/10.1186/1471-2458-12-594

